# Evaluation of the diagnostic value of YiDiXie™-SS, YiDiXie™-HS and YiDiXie™-D in endometrial carcinoma

**DOI:** 10.1101/2024.09.18.24313873

**Authors:** Yutong Wu, Yan Li, Chen Sun, Zhenjian Ge, Wenkang Chen, Yingqi Li, Shengjie Lin, Pengwu Zhang, Wuping Wang, Siwei Chen, Huimei Zhou, Xutai Li, Wei Li, Huiying Hu, Yanni Han, Yongqing Lai

## Abstract

**Background:** Endometrial cancer represents a significant challenge to human health and is associated with a considerable economic burden. Ultrasound is a commonly employed modality in the screening and preliminary diagnosis of endometrial cancer. However, false-positive uterine ultrasound results can lead to misdiagnosis and incorrect diagnostic curettage, while false-negative uterine ultrasound results can lead to missed diagnosis and delayed treatment. There is an urgent need to find convenient, cost-effective and non-invasive diagnostic methods to reduce the false-positive and false-negative rates of uterine ultrasound. The aim of the present study was to evaluate the diagnostic value of YiDiXie™-SS, YiDiXie™-HS and YiDiXie™-D in endometrial cancer.

**Patients and methods:** The final number of study subjects included in this study was 74, comprising 58 cases of malignant disease and 16 cases of benign disease. The remaining serum samples from the subjects were collected and tested using the YiDiXie™ all-cancer detection kit, with the objective of evaluating the sensitivity and specificity of the YiDiXie™-SS, YiDiXie™-HS and YiDiXie ™-D, respectively.

**Results:** The sensitivity of YiDiXie™-SS was 98.3% (95% CI: 90.9% - 99.9%) and its specificity was 62.5% (95% CI: 38.6% - 81.5%). This means that YiDiXie™-SS has an extremely high sensitivity and relatively high specificity in endometrial tumors.YiDiXie™-HS has a sensitivity of 82.8% (95% CI: 71.1% - 90.4%) and a specificity of 81.3% (95% CI: 57.0% - 93.4%). This means that YiDiXie™-HS has high sensitivity and specificity in endometrial tumors.YiDiXie™-D has a sensitivity of 70.7% (95% CI: 50.0% - 80.8%) and a specificity of 93.8% (95% CI: 71.7% - 99.7%). This means that YiDiXie™-D has relatively high sensitivity and very high specificity in endometrial tumors. The sensitivity of YiDiXie™-SS in patients with a positive uterine ultrasound was 95.0% (95% CI: 76.4% - 99.7%), while the specificity was 75.0% (95% CI: 30.1% - 98.7%). This indicates that the utilisation of YiDiXie ™-SS has the potential to reduce the incidence of false-positive uterine ultrasound results by 75.0% (95% CI: 30.1% - 98.7%), while simultaneously minimising the likelihood of malignancy leakage. The YiDiXie™-HS in patients with a negative uterine ultrasound was 81.6% (95% CI: 66.6% - 90.8%), while the specificity was 83.3% (95% CI: 55.2% - 97.0%). This suggests a reduction of 81.6% (95% CI: 66.6% - 90.8%) in the rate of false-negative uterine ultrasound with the application of YiDiXie™-HS. The sensitivity of YiDiXie™-D in uterine ultrasound positive patients was 75.0% (95% CI: 53.1% - 88.8%) and its specificity was 100% (95% CI: 51.0% - 100%). This means that YiDiXie™-D reduces the rate of false-positive uterine ultrasound by 100% (95% CI: 51.0% - 100%). YiDiXie™-D has a sensitivity of 68.4% (95% CI: 52.5% - 80.9%) and a specificity of 91.7% (95% CI: 64.6% - 99.6%) in patients with negative uterine ultrasound. This means that YiDiXie™-D reduces the false-negative rate of uterine ultrasound by 68.4% (95% CI: 52.5% - 80.9%) while maintaining high specificity.

**Conclusion:** YiDiXie™-SS has extremely high sensitivity and relatively high specificity in endometrial tumors.YiDiXie™-HS has high sensitivity and high specificity in endometrial tumors.YiDiXie™-D has relatively high sensitivity and extremely high specificity in endometrial tumors.YiDiXie™-SS has substantially reduced the rate of false positives on uterine ultrasound with essentially no increase in delayed treatment of endometrial cancer. YiDiXie ™ -HS significantly reduces the false negative rate of uterine ultrasound. YiDiXie™-D significantly reduces the false positive rate of uterine ultrasound or significantly reduces the false negative rate of uterine ultrasound while maintaining a high level of specificity. YiDiXie ™ tests are of significant diagnostic value in endometrial cancer, and are expected to solve the problem of the “high false positive rate” and “high false negative rate” of uterine ultrasound.

**Clinical trial number:** ChiCTR2200066840.

## INTRODUCTION

Endometrial cancer (EC) is one of the most prevalent malignant neoplasms in the field of obstetrics and gynaecology. Its prevalence is higher in developed countries where the prevalence of obesity is also higher^1^. The most recent data indicate that in 2022, there will be 420,242 new cases of endometrial cancer globally, representing 2.1% of the total number of new malignant tumours worldwide and ranking 15th^2^. Additionally, there will be 97,704 new deaths, accounting for 1% of the total number of deaths from malignant tumours globally and ranking 19th^2^. The incidence of endometrial cancer continues to increase on an annual basis, largely due to the prevalence of a high-fat diet and sedentary lifestyle in recent years. The treatment of endometrial cancer is based on a combination of surgical treatment, supplemented by radiotherapy and chemotherapy^3^. Studies have demonstrated that the five-year survival rate for stage I and II endometrial cancer can reach 95%, whereas the five-year survival rate for stage III and IV decreases to 60%^3-4^. Sheman EN^5^ and colleagues have developed a Relative Risk Index (RR) of endometrial cancer for patients with a body mass index (BMI) greater than 32, which is equal to 3.5. This indicates that patients are at a significantly increased risk of endometrial cancer. Consequently, endometrial cancer represents a significant health concern. Early screening has the dual benefit of improving survival outcomes and optimising survival rates. This is achieved by diagnosing and treating endometrial cancer at a curable stage. Furthermore, early screening reduces the burden and cost of treatment for patients.

Transvaginal ultrasonography (TVS) is a commonly employed diagnostic tool in the initial detection and screening of endometrial cancer^6^. It should be noted that transvaginal ultrasonography (TVS) has the potential to produce a significant number of false-positive results. A study by Smith-Bindman et al. revealed a 39% false-positive rate for transvaginal ultrasound measurements of endometrial thickness greater than 5 mm^7^. The occurrence of false-positive results has the potential to subject patients to a number of burdensome consequences. These include the necessity for invasive diagnostic procedures, such as curettage, which are costly and may cause distress. Additionally, patients may experience psychological distress, financial burden due to the costs of testing and treatment, and physical injury. It is therefore imperative to identify a convenient, cost-effective and non-invasive diagnostic method to reduce the rate of false-positive TVS.

Conversely, transvaginal ultrasound (TVS) has the potential to yield a considerable number of false-negative results. In a study conducted by Smith-Bindman and colleagues, which included 5,892 subjects, it was demonstrated that the false-negative rate of TVS for endometrial cancer surveillance is contingent upon the endometrial thickness criterion. When endometrial thickness was utilized as a criterion of 5 mm, the false-negative rate was approximately 5%. Additionally, the false-negative rate of TVS is influenced by age and the utilisation of tamoxifen medication, which results in an imprecise diagnosis of endometrial cancer^7^. In the event of a negative TVS result, patients are typically observed and followed up on a regular basis. A false negative TVS result indicates that a malignant tumour has been misdiagnosed as a benign condition, which may result in a delay in treatment, progression of the malignancy, and potentially even the development of an advanced stage. Consequently, patients will be forced to endure the detrimental consequences of an unfavourable prognosis, elevated treatment costs, diminished quality of life and diminished survival. It is therefore imperative to identify a convenient, cost-effective and non-invasive diagnostic method to reduce the false-negative rate of TVS.

The detection of novel tumour markers in serum has led to the identification of microRNAs as potential biomarkers for cancer. Shenzhen KeRuiDa Health Technology Co has developed an in vitro diagnostic test, the YiDiXie™ all-cancer tests (hereinafter referred to as ‘YiDiXie™ tests’), which are capable of detecting a variety of cancer types with a minimal amount of biological material, specifically 200 microlitres of whole blood or 100 microlitres of serum^8^. YiDiXie ™ tests comprises three distinct assays: YiDiXie™-HS, YiDiXie™-SS and YiDiXie™ -D.

The objective of this study is to assess the diagnostic efficacy of YiDiXie ™ -SS, YiDiXie ™ -HS and YiDiXie ™ -D in the context of endometrial cancer.

## PATIENTS AND METHODS

### Study design

This work constitutes a sub-study, entitled ‘Evaluating the value of YiDiXie ™ tests as an adjunctive diagnostic in a variety of tumours’, of the SZ-PILOT study (ChiCTR2200066840).

The SZ-PILOT study (ChiCTR2200066840) is a single-centre, prospective, observational study design. Subjects who had provided written, informed consent for the donation of residual samples at the time of admission or physical examination were enrolled in the study, and 0.5 ml of their residual serum samples were collected for the purposes of this study.

The study was conducted in a blinded manner. The laboratory personnel who performed YiDiXie™ tests and the Coretta laboratory technicians who determined the results of YiDiXie™ tests were both blinded to the subjects’ clinical information. The clinical experts who evaluated the clinical data of the subjects were also unaware of the results of YiDiXie™ tests.

The study was approved by the Ethics Committee of Peking University Shenzhen Hospital and was conducted in accordance with the International Conference on Harmonisation (ICH) Code of Practice for the Quality Management of Clinical Trials of Pharmaceuticals and the Declaration of Helsinki.

### Participants

The two groups of subjects were enrolled separately, and all subjects who met the inclusion criteria were consecutively included.

The study initially included inpatients with a suspected solid or haematological malignancy who had provided general informed consent for the donation of remaining samples. Individuals with a postoperative pathological diagnosis of malignant tumour were categorised within the malignant group, while those with a postoperative pathological diagnosis of benign disease were included in the benign group. Those subjects with ambiguous pathological findings were excluded from the study. A number of samples from the malignant group and samples from healthy subjects in the benign group have been used in previous studies^8^.

Subjects who failed the serum sample quality test prior to YiDiXie™ tests were excluded from the study. For further details regarding the inclusion and exclusion criteria, please refer to the previous article published by this research group^8^.

### Sample collection, processing

The serum samples utilized in this study were procured from surplus serum obtained from routine medical consultations, obviating the necessity for additional blood sampling.

Approximately 0.5ml of serum was collected from the remaining serum of subjects in the Medical Laboratory Department and stored at -80°C for subsequent utilisation in the YiDiXie™ tests.

### YiDiXie™ tests

YiDiXie ™ tests are conducted using the YiDiXie™ all-cancer detection kit, an in vitro diagnostic kit for use in fluorescent quantitative PCR instruments developed and manufactured by Shenzhen KeRuiDa Health Technology Co^8^. The test is an in vitro diagnostic kit designed for use in fluorescent quantitative PCR instruments. The test determines the presence of cancer in a subject’s body by measuring the expression levels of dozens of miRNA biomarkers in the serum^8^. The test predefines appropriate thresholds for each miRNA biomarker, thereby ensuring that each miRNA marker is highly specific^8^. Furthermore, it integrates these independent assays in a concurrent testing format, which significantly increases sensitivity while maintaining specificity for broad-spectrum cancers^8^.

YiDiXie™ tests are comprised of three distinct tests, each with unique characteristics. The YiDiXie™-HS, the YiDiXie™-SS and the YiDiXie™-D tests are characterised by high accuracy, precision and diagnostic capabilities, respectively^8^. The YiDiXie™-HS test has been developed with sensitivity and specificity as primary considerations^8^. The YiDiXie™-SS test has significantly expanded the number of miRNA tests, thereby achieving extremely high sensitivity for all malignancy types and all clinical stages^8^. The YiDiXie™-D test markedly elevates the diagnostic threshold of a singular miRNA examination, thereby attaining an exceptionally high sensitivity for all clinical stages of all malignant tumour types^8^. Furthermore, it achieves an exceptionally high specificity for the full range of malignant tumour types^8^.

YiDiXie ™ tests should be performed in accordance with the instructions provided in the YiDiXie™ Whole Cancer Test Kit. For further details on the procedure, please refer to the previous article in this series^8^.

The raw test results are subjected to analysis by the laboratory technicians of Shenzhen KeRuiDa Health Technology Co., Ltd., who determine the results of YiDiXie ™ tests as either ‘positive’ or ‘negative’^8^.

### Ultrasound diagnosis

The diagnostic findings of the ultrasound of the uterus are classified as either positive or negative. A positive diagnosis is indicated when the findings are indicative of a malignant tumour, or when the probability of a malignant tumour is higher than that of a benign tumour. In the event that the diagnosis is positive, more certain, or indicative of a benign tumour, or if the diagnosis is ambiguous, it will be classified as negative.

### Extraction of clinical data

The data set comprised clinical, pathological, laboratory and imaging data extracted from the inpatient medical records or physical examination reports of the subjects. Clinical staging was conducted by trained clinicians in accordance with the AJCC staging manual (seventh or eighth edition)^9-10^.

### Statistical analyses

Descriptive statistics were employed to present the data pertaining to the demographic and baseline characteristics. For categorical variables, the number and percentage of subjects in each category were calculated. For continuous variables, the total number of subjects (n), mean, standard deviation (SD) or standard error (SE), median, first quartile (Q1), third quartile (Q3), minimum, and maximum values were calculated. The 95% confidence intervals (CIs) for the multiple indicators were calculated using the Wilson (score) method.

## RESULTS

### Participant disposition

A total of 74 study subjects were ultimately included in this investigation, comprising 58 cases belonging to the malignant group and 16 cases belonging to the benign group. The demographic and clinical characteristics of the 74 study subjects are presented in Table 1 for reference.

**Table 1.**
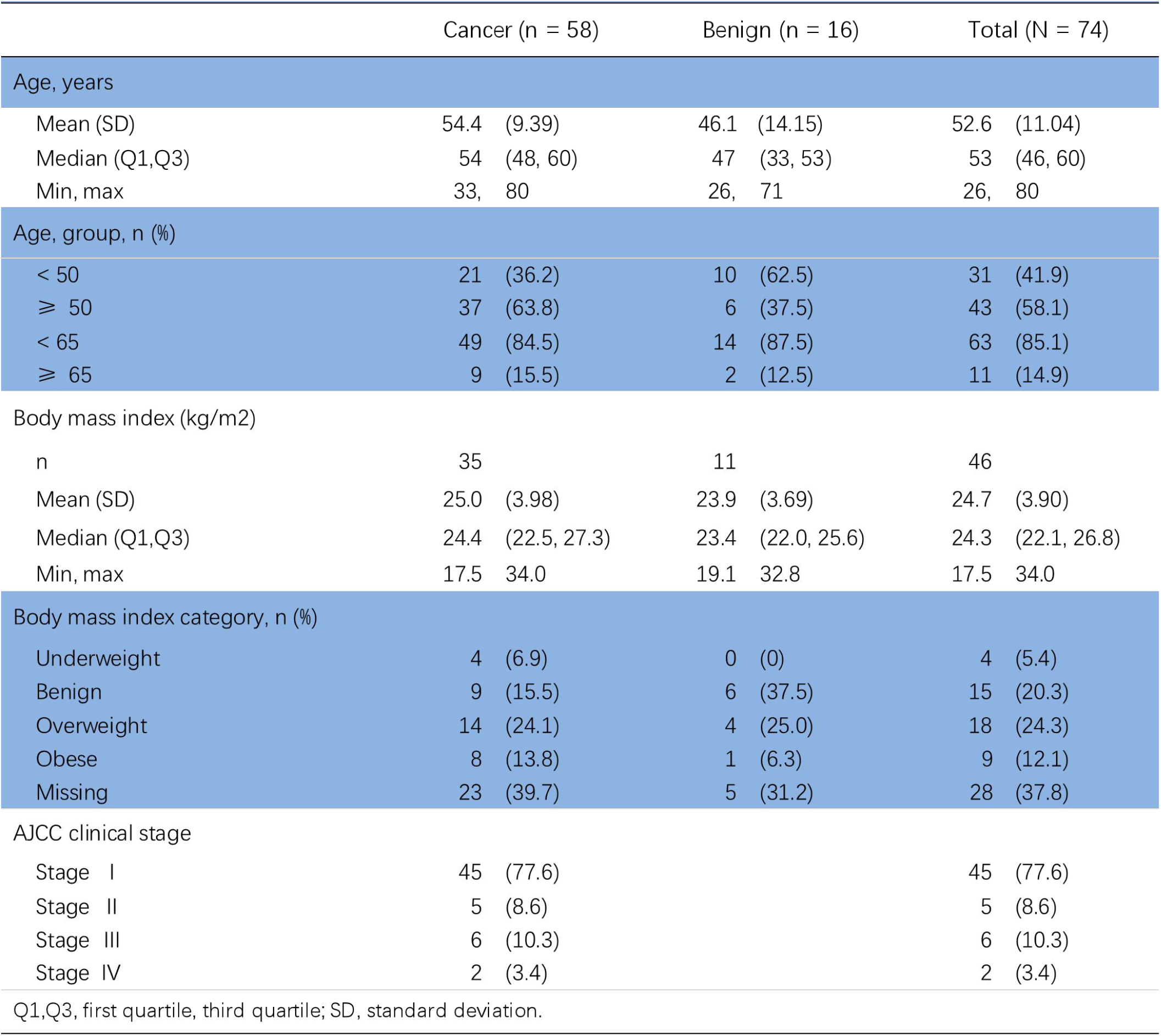
Participants’ demographic and clinical manifestation.

|  | Cancer (n = 58) | Benign (n = 16) | Total (N = 74) |
| --- | --- | --- | --- |
| Age, years |  |  |  |
| Mean (SD) | 54.4 (9.39) | 46.1 (14.15) | 52.6 (11.04) |
| Median (Q1,Q3) | 54 (48, 60) | 47 (33, 53) | 53 (46, 60) |
| Min, max | 33, 80 | 26, 71 | 26, 80 |
| Age, group, n (%) |  |  |  |
| < 50 | 21 (36.2) | 10 (62.5) | 31 (41.9) |
| ≥ 50 | 37 (63.8) | 6 (37.5) | 43 (58.1) |
| < 65 | 49 (84.5) | 14 (87.5) | 63 (85.1) |
| ≥ 65 | 9 (15.5) | 2 (12.5) | 11 (14.9) |
| Body mass index (kg/m<sup>2</sup>) |  |  |  |
| n | 35 | 11 | 46 |
| Mean (SD) | 25.0 (3.98) | 23.9 (3.69) | 24.7 (3.90) |
| Median (Q1,Q3) | 24.4 (22.5, 27.3) | 23.4 (22.0, 25.6) | 24.3 (22.1, 26.8) |
| Min, max | 17.5 34.0 | 19.1 32.8 | 17.5 34.0 |
| Body mass index category, n (%) |  |  |  |
| Underweight | 4 (6.9) | 0 (0) | 4 (5.4) |
| Benign | 9 (15.5) | 6 (37.5) | 15 (20.3) |
| Overweight | 14 (24.1) | 4 (25.0) | 18 (24.3) |
| Obese | 8 (13.8) | 1 (6.3) | 9 (12.1) |
| Missing | 23 (39.7) | 5 (31.2) | 28 (37.8) |
| AJCC clinical stage |  |  |  |
| Stage I | 45 (77.6) |  | 45 (77.6) |
| Stage II | 5 (8.6) |  | 5 (8.6) |
| Stage III | 6 (10.3) |  | 6 (10.3) |
| Stage IV | 2 (3.4) |  | 2 (3.4) |

Q1,Q3, first quartile, third quartile; SD, standard deviation.

The two groups of study subjects were found to be comparable in terms of demographic and clinical characteristics (see Table 1). The mean age of the subjects was 52.6 years (standard deviation 11.04).

### Diagnostic performance of YiDiXie™-SS

As shown in Table 2, the sensitivity of YiDiXie™ -SS was 98.3% (95% CI: 90.9% - 99.9%) and its specificity was 62.5% (95% CI: 38.6% - 81.5%) 61.2% (49.2% - 72.0%). This means that YiDiXie ™ -SS has extremely high sensitivity and relatively high specificity in endometrial tumors.

**Table 2.** The performance of YiDiXie™-SS.

|  | Cancer | Benign | Total |
| --- | --- | --- | --- |
|  | 58 | 16 | 74 |
| Positive | 57 | 6 | 63 |
| Negative | 1 | 10 | 11 |

SEN = 57/58 = 98.3% (90.9% - 99.9%)
SPE = 10/16 = 62.5% (38.6% - 81.5%)
SEN, Sensitivity. SPE, Specificity. Two-sided 95% Wilson confidence intervals were calculated.

### Diagnostic performance of YiDiXie™-HS

As shown in Table 3, the sensitivity of YiDiXie™ -HS was 82.8% (95% CI: 71.1% - 90.4%) and its specificity was 81.3% (95% CI: 57.0% - 93.4%).This means that YiDiXie ™ -HS has high sensitivity and specificity in endometrial tumors.

**Table 3.** The performance of YiDiXie™-HS.

|  | Cancer | Benign | Total |
| --- | --- | --- | --- |
|  | 58 | 16 | 74 |
| Positive | 48 | 3 | 51 |
| Negative | 10 | 13 | 23 |

SEN = 48/58 = 82.8% (71.1% - 90.4%)
SPE = 13/16 = 81.3% (57.0% - 93.4%)
SEN, Sensitivity. SPE, Specificity. Two-sided 95% Wilson confidence intervals were calculated.

### Diagnostic performance of YiDiXie™-D

As shown in Table 4, the sensitivity of YiDiXie™ -D was 70.7% (95% CI: 50.0% - 80.8%) and its specificity was 93.8% (95% CI: 71.7% - 99.7%). This means that YiDiXie ™ -D has relatively high sensitivity and very high specificity in endometrial tumors.

**Table 4.**
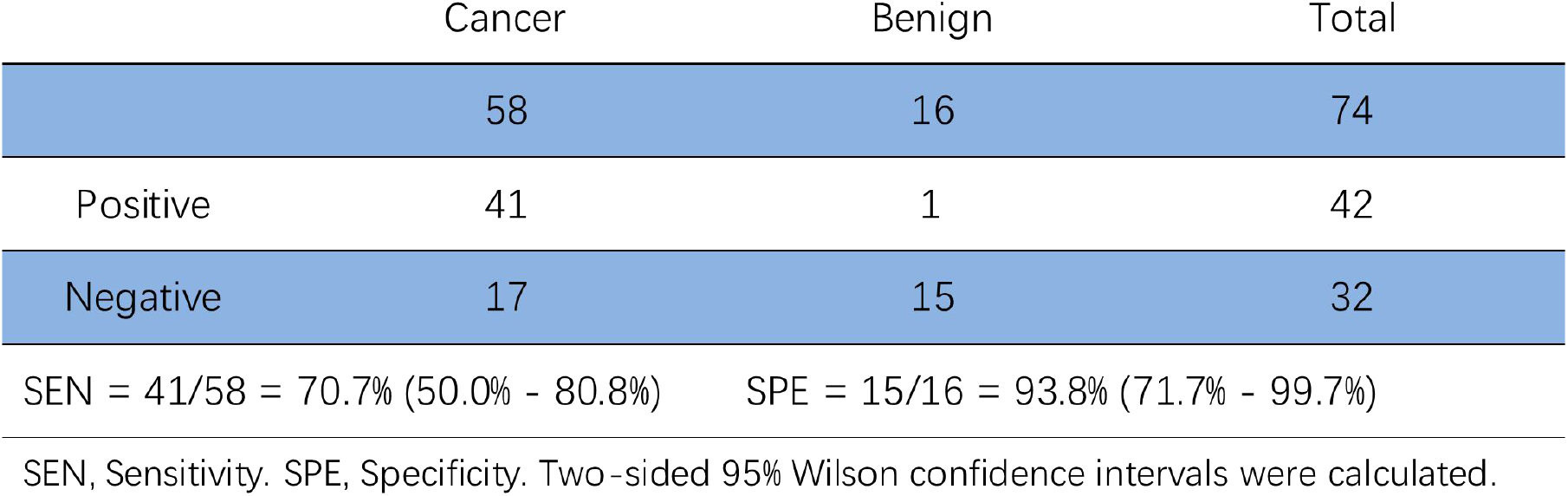
The performance of YiDiXie™-D.

|  | Cancer | Benign | Total |
| --- | --- | --- | --- |
|  | 58 | 16 | 74 |
| Positive | 41 | 1 | 42 |
| Negative | 17 | 15 | 32 |

SEN, Sensitivity. SPE, Specificity. Two-sided 95% Wilson confidence intervals were calculated.

### Diagnostic performance of ultrasound

As shown in Table 5, the uterine ultrasound sensitivity was 34.5% (95% CI: 23.6% - 47.3%) and its specificity was 75.0% (95% CI: 50.5% - 89.8%).

**Table 5.**
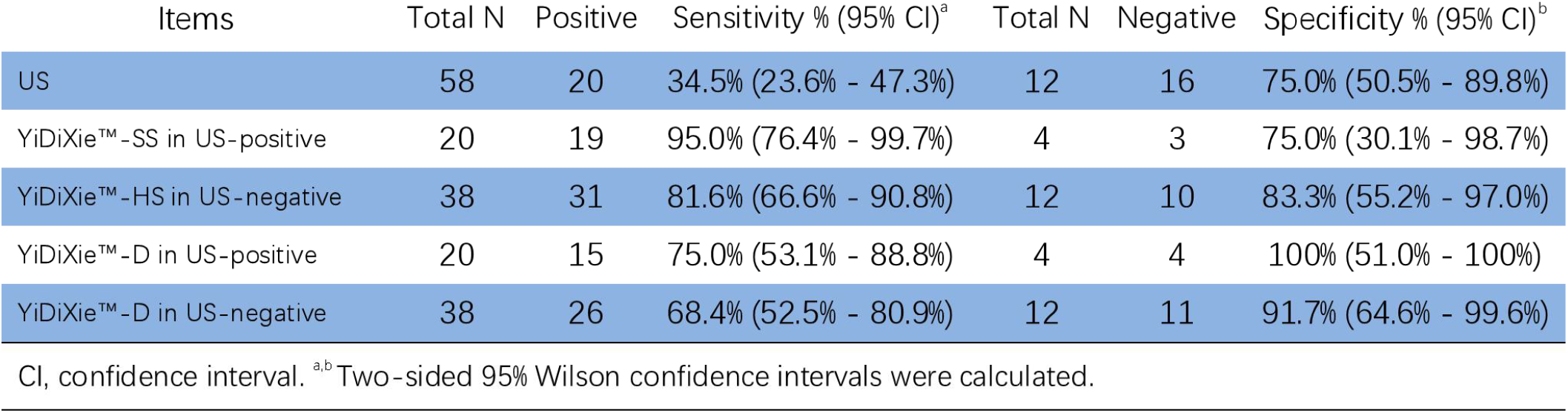
Performance of different Items.

### Diagnostic performance of YiDiXie™-SS in uterine ultrasound-positive patients

In order to address the issue of a high false-positive rate associated with uterine ultrasound, YiDiXie™-SS was employed in the context of patients with a positive uterine ultrasound result.

As illustrated in Table 5, the sensitivity of YiDiXie™-SS in uterine ultrasound-positive patients was 95.0% (95% CI: 76.4% - 99.7%), while the specificity was 75.0% (95% CI: 30.1% - 98.7%). This indicates that the utilisation of YiDiXie™-SS has the potential to reduce the incidence of false-positive uterine ultrasound results by 75.0% (95% CI: 30.1% - 98.7%) while simultaneously minimising the risk of malignant tumour leakage.

### The Diagnostic performance of YiDiXie™-HS in uterine ultrasound-negative patients

In order to address the challenge of the high false-negative rate of uterine ultrasound, the application of YiDiXie™-HS was conducted in patients who had undergone a negative uterine ultrasound.

As demonstrated in Table 5, the sensitivity of YiDiXie™-HS in patients with negative uterine ultrasound results was 81.6% (95% CI: 66.6% - 90.8%), while the specificity was 83.3% (95% CI: 55.2% - 97.0%).

This indicates a reduction of 81.6% (95% CI: 66.6% - 90.8%) in the incidence of false-negative uterine ultrasound results with the implemen tation of YiDiXie™-HS.

### Diagnostic performance of YiDiXie™-D in uterine ultrasound-positive patients

To further minimize the rate of uterine ultrasound false positives, YiDiXie™-D, which has a relatively high sensitivity and very high specificity, was therefore applied.

As shown in Table 5, YiDiXie ™ -D had a sensitivity of 75.0% (95% CI: 53.1% - 88.8%) and its specificity was 100% (95% CI: 51.0% - 100%) in patients with positive uterine ultrasound. This means that YiDiXie ™-D reduces the false positive rate of uterine ultrasound by 100% (95% CI: 51.0% - 100%).

### The Diagnostic performance of YiDiXie™-D in uterine ultrasound-negative patients

In order to reduce the false-negative rate of uterine ultrasound while maintaining a high level of specificity, YiDiXie™-D, which has a relatively high sensitivity and very high specificity, was applied.

As shown in Table 5, YiDiXie ™ -D had a sensitivity of 68.4% (95% CI: 52.5% - 80.9%) and its specificity was 91.7% (95% CI: 64.6% - 99.6%) in patients with negative uterine ultrasound. This means that YiDiXie ™ -D reduces the rate of false-negative uterine ultrasound by 68.4% (95% CI: 52.5% - 80.9%) while maintaining high specificity.

## DISCUSSION

### Clinical significance of YiDiXie™-SS in uterine ultrasound-positive patients

In patients with a positive uterine ultrasound, the utilisation of supplementary diagnostic techniques is crucial for optimising sensitivity and specificity. In general, a positive result on a uterine ultrasound scan is followed by diagnostic curettage rather than radical surgery. Consequently, a false-positive uterine ultrasound does not result in significant adverse outcomes such as major surgical trauma, organ removal, or loss of function. Therefore, the probability of an undiagnosed malignant tumour is considerably higher than that of a misdiagnosed benign tumour in patients with a positive uterine ultrasound. Accordingly, YiDiXie ™ -SS was selected to diminish the false-positive rate of uterine ultrasound, exhibiting an exceptionally high sensitivity but a somewhat diminished specificity.

As demonstrated in Table 5, the YiDiXie™-SS in patients with a positive uterine ultrasound was 95.0% (95% CI: 76. The sensitivity was 95.0% (95% CI: 76.0% - 99.7%), while the specificity was 75.0% (95% CI: 30.1% - 98.7%). These results indicate that, while maintaining a sensitivity of nearly 100%, YiDiXie ™ -SS reduced the number of false positives identified by uterine ultrasound by 75.0% (95% CI: 30.1% - 98.7%).

The results demonstrate that YiDiXie ™ -SS markedly diminishes the likelihood of erroneous diagnostic curettage in patients with benign endometrial disease, while concurrently exhibiting minimal propensity for unidentified malignancies. Consequently, YiDiXie™-SS is an effective solution that addresses a significant clinical need and has important clinical significance and wide application prospects.

### Clinical implications of YiDiXie™-HS in uterine ultrasound-negative patients

In patients with a negative uterine ultrasound, the sensitivity and specificity of subsequent diagnostic techniques are of significant importance. A higher false-negative rate results in an increased number of malignant tumours being under-diagnosed. This can lead to delays in treatment. A higher false-positive rate indicates an increased likelihood of misdiagnosis of benign diseases Accordingly, the highly sensitive and specific YiDiXie ™ -HS was selected as a means of reducing the false negative rate of uterine ultrasound.

As demonstrated in Table 5, the sensitivity of YiDiXie ™ -HS in patients with negative uterine ultrasound was 81.6% (95% CI: 66.6% - 90.8%), while the specificity was 83.3% (95% CI: 55.2% - 97.0%). These findings demonstrate that YiDiXie™-HS has the potential to reduce false-negative uterine ultrasound results by 81.6% (95% CI: 66.6% - 90.8%).

The aforementioned results indicate that YiDiXie™-HS markedly diminishes the likelihood of false-negative uterine ultrasound results in cases where malignancies are overlooked. It can therefore be concluded that YiDiXie ™ -HS meets the clinical requirements and has significant clinical importance and potential for wide application.

### Clinical implications of YiDiXie™-D in endometrial tumors

In patients with endometrial tumors, YiDiXie™ -D, with its relatively high sensitivity and very high specificity, can be used to further reduce the rate of false-positive uterine ultrasound or to significantly reduce its false-negative rate while maintaining a high level of specificity.

As shown in Table 5, the sensitivity of YiDiXie™ -D in uterine ultrasound-positive patients was 75.0% (95% CI: 53.1% - 88.8%), and its specificity was 100% (95% CI: 51.0% - 100%); the sensitivity of YiDiXie™-D in uterine ultrasound-negative patients was 68.4% (95% CI: 52.5% - 80.9%) and its specificity was 91.7% (95% CI: 64.6% - 99.6%). These results indicate that YiDiXie™-D reduces false positives by 100% (95% CI: 51.0% - 100%) or reduces false negatives by 68.4% (95% CI: 52.5% - 80.9%) while maintaining a high specificity, respectively.

The above results imply that YiDiXie ™ -D further reduces the risk of wrong diagnostic curettage for endometrial tumors. Therefore, YiDiXie™-D well meets the clinical needs and has important clinical significance and wide application prospects.

### YiDiXie™ tests can address the two challenges of endometrial cancer

Firstly, the use of YiDiXie ™ -SS has the potential to significantly reduce the administrative workload of obstetricians and gynaecologists, thereby facilitating the timely diagnosis and prompt treatment of malignancy cases that would otherwise be delayed. In the event of a positive result on the uterine ultrasound, a diagnostic curettage is typically performed on the patient. The timely completion of these diagnostic scrapings is contingent upon the number of obstetricians and gynaecologists available. In numerous regions across the globe, appointments are scheduled months or even years in advance. This inevitably results in delays to the treatment of malignant cases, with the consequence that patients with positive ultrasound results awaiting diagnostic curettage may show evidence of disease progression or even distant metastases. As demonstrated by the results, YiDiXie ™ -SS was observed to reduce the rate of false-positive uterine ultrasound by 75.0% (95% CI: 30.1% - 98.7%), with essentially no increase in missed malignancies. Consequently, YiDiXie ™ -SS has the potential to markedly reduce the superfluous workload of obstetricians and gynaecologists, thereby facilitating the prompt diagnosis and treatment of endometrial cancer and other conditions that would otherwise be delayed.

Secondly, YiDiXie ™ -HS has the potential to significantly reduce the risk of underdiagnosis of endometrial cancer. In the event of a negative uterine ultrasound, the likelihood of endometrial cancer is typically discounted for the time being. The high rate of false-negative uterine ultrasound results consequently results in delayed treatment for a considerable number of endometrial cancer patients. As demonstrated in the results, YiDiXie™ -HS was observed to reduce the false-negative rate of uterine ultrasound by 81.6% (95% CI: 66.6% - 90.8%). Consequently, YiDiXie ™ -HS markedly diminishes the likelihood of false-negative uterine ultrasound results for undiagnosed malignancies, thereby expediting the diagnosis and treatment of patients with endometrial cancer who would otherwise experience delays in treatment.

Again, YiDiXie ™ -D is expected to further address the challenges of “high false positive rate” and “high false negative rate”. As shown in Table 5, YiDiXie ™ -D reduced the false-positive rate of uterine ultrasound by 100% (95% CI: 51.0% - 100%), or 68.4% (95% CI: 52.5% - 80.9%) while maintaining high specificity. Thus, YiDiXie™-D further reduced the risk of false diagnostic curettage for endometrial tumors.

Finally, YiDiXie ™ tests enables the prompt identification of endometrial cancer. From the patient’s perspective, the YiDiXi™ test requires only a minimal quantity of blood, enabling the diagnostic process to be completed without the need for invasive procedures and without the patient having to leave their home. A mere 20 microlitres of serum is necessary to complete a YiDiXie™ test, which is equivalent to approximately one drop of whole blood (one drop of whole blood is approximately 50 microlitres, yielding 20-25 microlitres of serum). In consideration of the pre-test sample quality assessment test and two or three repetitions, a volume of 0.2 ml of whole blood is sufficient for YiDiXie™ tests. The 0.2 ml of finger blood can be collected at home by the average patient using a finger blood collection needle, obviating the need for venous blood collection by medical personnel. This allows the patient to complete the diagnostic process non-invasively without having to leave their home.

Conversely, the diagnostic capacity of YiDiXie ™ tests are nearly unlimited. Figure 1 illustrates the fundamental operational sequence of YiDiXie ™ tests, demonstrating that the test does not necessitate the involvement of a physician or specialized medical equipment, nor does it require the assistance of medical personnel for blood collection. Consequently, YiDiXie ™ tests are entirely independent of the number of medical personnel and medical institutions, and its testing capacity is effectively limitless. Consequently, YiDiXie ™ tests facilitates prompt diagnosis of endometrial cancer, obviating the need for patients to await appointments with trepidation.

**Figure 1.**
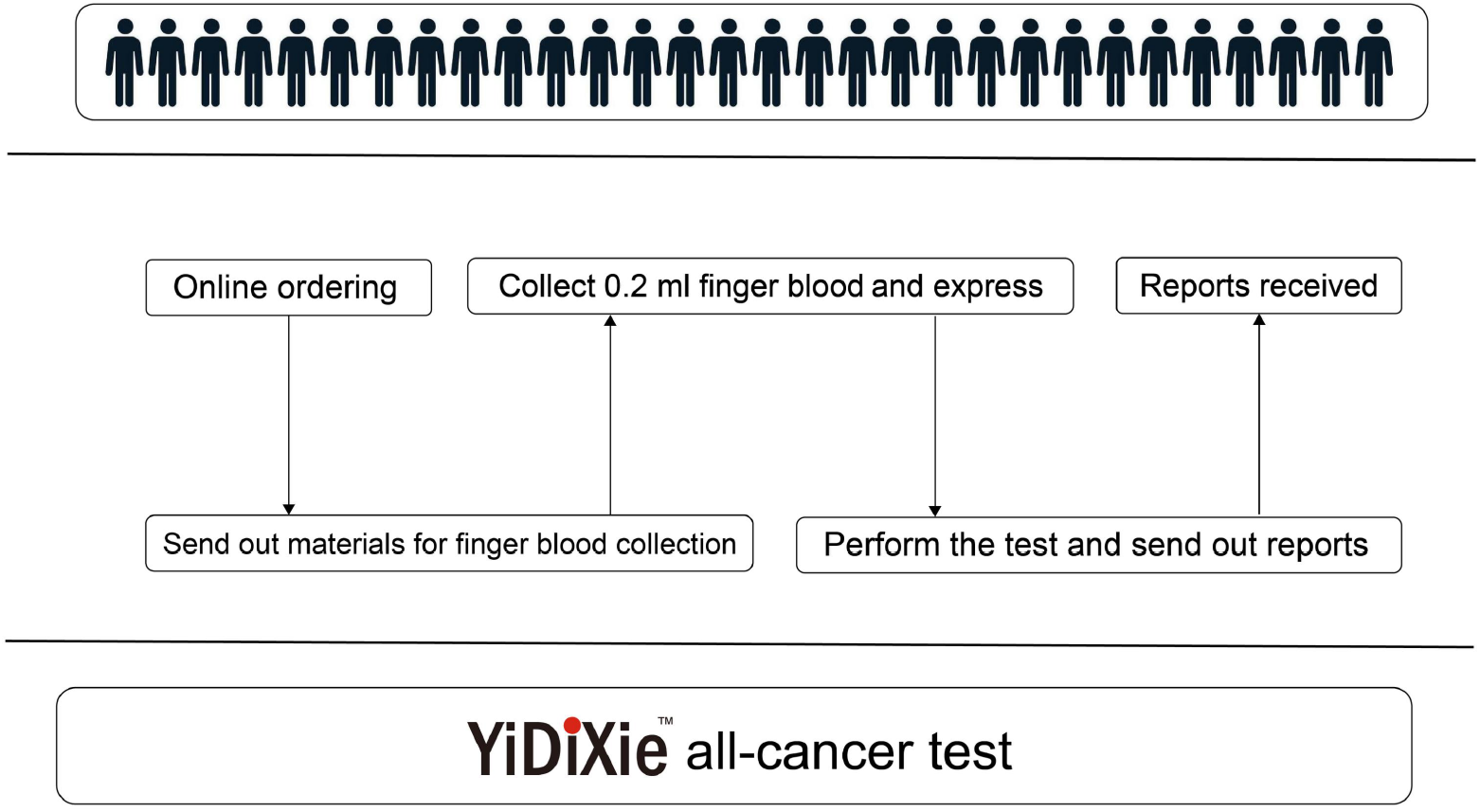
Basic flowchart of YiDiXie™ tests.

In summary, YiDiXie ™ tests are a valuable diagnostic tool for endometrial cancer. It is anticipated that YiDiXie ™ tests will address the issues of a high false-positive rate and a high false-negative rate in endometrial cancer.

### Limitations of the study

Firstly, the number of cases included in this study was relatively small, and it would be beneficial for future clinical studies to include larger sample sizes in order to facilitate further assessment.

Secondly, this study was a case-control study of malignant and benign tumours in inpatients, and further assessment would benefit from cohort studies in the natural population of endometrial cancer.

Finally, it should be noted that this study was conducted at a single centre, which may have introduced some degree of bias into the results. Further assessment would benefit from the inclusion of multicentre studies.

## CONCLUSION

YiDiXie™-SS has extremely high sensitivity and relatively high specificity in endometrial tumors.YiDiXie ™-HS has high sensitivity and high specificity in endometrial tumors.YiDiXie ™ -D has relatively high sensitivity and extremely high specificity in endometrial tumors.YiDiXie ™ -SS has substantially reduced the rate of false positives on uterine ultrasound with essentially no increase in delayed treatment of endometrial cancer. YiDiXie™ -HS significantly reduces the false negative rate of uterine ultrasound. YiDiXie ™ -D significantly reduces the false positive rate of uterine ultrasound or significantly reduces the false negative rate of uterine ultrasound while maintaining a high level of specificity. YiDiXie ™ tests are of significant diagnostic value in endometrial cancer, and are expected to solve the problem of the “high false positive rate” and “high false negative rate” of uterine ultrasound.

## Data Availability

All data produced in the present study are contained in the manuscript.

